# Genome-Wide Association Study of Estradiol Levels, and the Causal Effect of Estradiol on Bone Mineral Density

**DOI:** 10.1101/2021.07.01.21259826

**Authors:** Daniel Schmitz, Weronica E. Ek, Elin Berggren, Julia Höglund, Torgny Karlsson, Åsa Johansson

## Abstract

**Context:** Estradiol is the primary female sex hormone and plays an important role for skeletal health in both sexes. Several enzymes are involved in estradiol metabolism but few genome-wide association studies (GWAS) have been performed to characterize the genetic contribution to variation in estrogen levels.

**Objective:** Identify genetic loci affecting estradiol levels and estimate causal effect of estradiol on bone mineral density (BMD).

**Design:** We performed GWAS for estradiol in males (N = 147,690) and females (N = 163,985) from UK Biobank (UKB). Estradiol was analyzed as a binary phenotype; above/below detection limit (175 pmol/L). We further estimated the causal effect of estradiol on BMD using Mendelian randomization.

**Results:** We identified 14 independent loci associated (P<5×10^−8^) with estradiol levels in males, of which one (*CYP3A7*) was genome-wide and seven nominally (P<0.05) significant in females. In addition, one female specific locus was identified. Most loci contain functionally relevant genes that have not been discussed in relation to estradiol levels in previous GWAS. For example, *SRD5A2*, which encodes a steroid 5-alpha reductase that is involved in processing androgens, and *UGT3A1* and *UGT2B7* which encode enzymes likely to be involved in estradiol elimination. The allele that tags the O blood group, at the *ABO* locus, was associated with higher estradiol levels. We identified a causal effect of high estradiol levels on increased BMD in both males (P=1.58×10^−11^) and females (P=7.48×10^−6^).

**Conclusion:** Our findings further support the importance of the body’s own estrogen to maintain skeletal health in males and in females.

## Introduction

Estrogen, the primary female sex hormone, is responsible for the development of the female reproductive system and secondary sex characteristics. Estrogen acts primarily as a growth hormone for the reproductive organs and regulates the menstrual cycle in women^1^. Additionally, it plays a critical role in male sexual function^2^. There are three major forms of estrogen: estrone, estradiol and estriol. Estradiol is the major and most potent form^3^. In females, high estrogen levels have been associated with deep vein thrombosis^4^ and increased risk for certain cancers, including breast cancer^5^ in women. In males, higher levels have been suggested to be associated with reduced risk of type 2 diabetes (T2D)^6^. Estrogen is also an important regulator of bone metabolism in both females and males^7^. When estrogen levels drop during menopause, the cells that synthesize bone (osteoblasts) are unable to effectively produce bone mass^8^. The loss of ovarian estrogens after menopause has been associated with reduced bone mineral density (BMD) and higher risk of osteoporosis^9,10^.

Previously published genome-wide association studies (GWAS) for estrogen have been performed in cohorts of up to 11,000 individuals, mostly of European descent, both male and female and stratified by sex^11–15^. In addition, a GWAS was recently performed for sex hormone levels in UK biobank (UKB), a study which mainly focused on testosterone^16^. They identified strong sex-specific genetic effects on serum testosterone levels, but did not consider the results from estradiol in women, due to the strong link between estradiol levels and menopausal status/age of menopause. Previous studies have also addressed the causal effect of estradiol and testosterone levels in relation to disease risk using Mendelian randomization (MR)^15–17^. MR uses genetic data to evaluate a potential causal relationship between an exposure, for example hormones, and disease risk. The underlying assumption in MR is that if a hormone is involved in a disease process, then the genetic factors influencing the levels of the hormone should only affect disease risk through their effects on hormone levels, and not through any alternative pathways. MR is conceptually similar to randomized controlled trials because genotypes are assigned randomly already at conception. MR can therefore be used to overcome some of the limitations of observational studies, such as bias due to unmeasured or unknown confounding. Likewise, reverse causation is also avoided due to genotypes being assigned before the disease manifests. For testosterone, many independent genetic variants were identified and included in the previous MR. They concluded that testosterone increases the risk for polycystic ovary syndrome (PCOS) and type 2 diabetes in women but decreases the risk for type 2 diabetes in men^16^. Even though the number of GWAS significant findings for estradiol has been quite limited in previous studies^11–15^, those variants have been used in MR studies to address the causal effect of estradiol on disease risk. Two recent MR studies, using five and two SNPs respectively, identified a beneficial effect of estradiol on bone mineral density (BMD) in males^15,17^, but no study has yet been performed in females.

By using the largest cohort with estradiol levels measured; almost half a million participants, we have been able to further highlight novel genes, important for regulating estradiol levels. Most assays for measuring estradiol levels are not very sensitive, and the methods used in UKB had a detection limit of 175 pmol/L. Normal ranges of serum estradiol levels are 36.7-183.6 pmol/L in men, 73.4 – 2753.5 pmol/L in premenopausal women and 0 – 73.4 pmol/L in postmenopausal women^18^. This results in a large fraction of the samples, especially men and postmenopausal women, having levels below detection limit. Similar to a previous GWAS for estradiol^16^, we have therefore analyzed estradiol as a binary phenotype (above or below detection limit), instead of removing all individuals below detection limit. We have stratified the cohort by sex and menopausal status, to enable comparing genetic effects across strata. Thanks to the larger sample size in UKB compared to previous studies, we were able to identify enough genetic variants associated with estradiol levels to permit causal effect estimation of estradiol on bone mineral density, through MR analysis, also in females.

## Material and Methods

### Sample

UKB is a cross sectional cohort of about 500,000 participants, born between 1939 and 1970, that were recruited 2006—2010. Detailed information was collected on lifestyle and anthropometric traits, and on female specific factors, that are likely to effect hormone levels, including use of birth control pills, hormone replacement treatments, pregnancies and menopause. Participants also provided blood samples that were used to genotype approximately 800,000 single nucleotide polymorphisms (SNPs) across the genome and to measure levels of estradiol. The application to use data from UKB has been approved (under project number 41143). UKB has an ethics permit from the National Research Ethics Committee (REC reference 11 / NW / 0382). Phenotype data for this project was extracted from the UKB database April 2019. All analyses performed in this study are based on the samples and information collected at the participants initial visit to the assessment center. Ethical approval for the analyses performed in this study has also been approved by the Swedish Ethical Review Authority (Dnr: 2020-04415)

### Genotype data

A total of 438,417 participants had been genotyped using the UKB Axiom array, and another 49,994 participants had been genotyped using the similar (95% common marker content) UK BiLEVE array. SNPs had been imputed using UK10K and 1000 genomes phase 3, as reference panels^19,20^. For this project we used imputed data from the third release which contained a total of 93,093,070 SNPs. We only analyzed SNPs with minor allele frequency (MAF) > 0.01, and removed SNPs deviating from Hardy-Weinberg (P-value < 1×10^−20^) and markers with more than 5% missing genotype data, and with imputation quality < 0.3. After quality control (QC), a total of 7,651,231 autosomal SNPs, and 220,486 SNPs on the X-chromosome remained. We included Caucasian participants clustering with regards to their genetic principal components. We also excluded first- and second-degree relatives (genetic relationship > 0.044), using kinship data. Samples with sex discordance, high heterozygosity/missingness, and with more than 5% missing genotypes were also excluded. After QC and exclusion, 361,975 unrelated Caucasian participants remained (167,168 males and 194,807 females).

### Estradiol measurements

Estradiol had been measured by two step competitive analysis on a Beckman Coulter Unicel Dxl 800. We only included measurements from the blood samples that were collected during the participants initial visit to the assessment center. Among all participants, only 76,668 had estradiol measurements above detection limit (175 pmol/L). As the majority of the participants had measurements below detection limit, estradiol levels were analyzed as a binary phenotype (above/below detection limit).

### GWAS and sensitivity analyses

GWAS was performed in males and females separately, using logistic regression with an additive genetic model implemented in PLINK version 2. The following covariates were included in both male and female analyses: age, body mass index (BMI), the first ten genetic principal components (to adjust for population structures and ethnic origins), and a binary indicator variable for UKB Axiom versus UK BiLEVE genotyping array, to adjust for any array differences. For females, hormone-replacement therapy (HRT) (current, ever or never), oral contraceptive use (current, ever or never), number of live births, menopausal status and whether they have undergone hysterectomy, were also included. Women who were unsure about menopausal status were excluded. For this study, we applied a P-value threshold of 5×10^−8^, which is commonly used in GWAS when using a MAF cutoff of 1%^21^, and corresponds to a correction of 1 million independent tests (SNPs). Manhattan plots were generated using the R package hudson. We applied the clump-function in PLINK 1.90 to define start and end positions for each locus with the main parameters that determines the clumping being set to: *R*^2^=0.1, *p*_1_=5×10^−8^ and *p*_2_=1×10^−4^. We subsequently performed conditional analyses for each locus, conditioning on the SNP with the lowest P-value from each locus. If there was any significant conditional SNP, this was repeated by also conditioning on the conditional SNP from the previous analyses, until no additional significant associations remained.

Several sensitivity analyses were performed for the GWAS. First, we stratified females into being pre- and postmenopausal. Secondly, we excluded all participants with a cancer diagnosis prior to assessment (when blood samples were taken). Finally, we also analyzed males and females with testosterone and sex hormone-binding globulin (SHBG) levels included as covariates, in addition to the other covariates and with age at menopause as a covariate in the female postmenopausal stratum.

### Annotation of GWAS lead SNPs

To identify likely target genes for the associated variants, we first identified genes containing the lead SNPs, or the closest gene(s) for each individual lead SNP. We also used HaploReg v4.1^22^ to identify potential functional effects by the individual lead SNP, or any variant in linkage disequilibrium (LD, *R*^2^>0.8) with the lead SNP. We then checked for overlap between the lead SNPs and expression quantitative trait loci (eQTL) using data from the Genotype-Tissue Expression (GTEx) project^23^. We considered our lead SNP to overlap with an eQTL if our lead SNP was in LD (*R*^2^>0.8) with a lead SNP for an eQTL. To identify overlap with other phenotypes from previously published GWAS we downloaded summary statistics from the GWAS catalog (Version downloaded: 1.0.3, date: 2020-11-13). We defined signals to be overlapping if a lead SNP from a previous GWAS was in LD (*R*^2^>0.8) with any of our lead SNPs.

### Mendelian Randomization

Firstly, we undertook a one-sample MR approach to investigate the causal effect of estradiol on BMD in UKB, in males and females separately. BMD was measured as heel BMD, based on an ultrasound measurement of the calcaneus (heel bone), using the Sahara Clinical Bone Sonometer. We used T-score, automated (data-field 78 in UKB). The T-score is the number of standard deviations a person’s BMD differs from the mean BMD of their respective sex. The main MR analyses were performed with the R package gsmr (version 1.0.8)^24^. The GSMR approach allows both for heteroskedastic and correlated data via the variance-covariance matrix, and identifies and removes pleiotropic outliers prior to causal effect estimation. The default LD threshold was used (*R*^2^=0.05) and the significance threshold for an SNP being identified as an outlier, and therefore not belonging to the set of valid instruments, was set to the default value of *a*=0.01. GSMR assumes a fixed effects model with the test statistic following a *χ*^2^ distribution with 1 degree of freedom. We further performed sensitivity analyses, applying inverse variance weighted, weighted median, and the MR-Egger method, using the “MendelianRandomization” tool implemented in R. The MR-Egger intercept test was used to assess the possible presence of directional pleiotropy in the set of genetic instruments, where a deviation from zero intercept indicates pleiotropy^25^. Both the inverse variance weighted and the MR-Egger methods assume a random effects model, with underdispersion not being allowed. The standard error of the estimate for the weighted median method is estimated using bootstrapping. If heterogeneity is present in the data, these methods would tend to show higher p-values as compared GSMR, as GSMR is expected to remove outliers. Secondly, the MR results were further replicated by two-sample MR, using the GWAS summary statistics for BMD from the GEFOS study^26^. This was done in order to avoid any weak-instrument bias towards the confounded association, potentially present in one-sample MR analyses. We used GWAS summary statistics for lumbar spine BMD (LSBMD) from the 2012 data release on www.gefos.org (downloaded: 2020-11-13), which was the only dataset with summary statistics for males and females separately that did not include samples from UKB. The two sets (for males and females) of genetic variants that had been selected in UKB were tested independently using GEFOS BMD summary statistics for males and females respectively. The overlap, with regards to SNPs, was not perfect between UKB and GEFOS. We therefore revised the set of SNPs for the MR in GEFOS slightly. For each of our GWAS locus, we selected the most significant SNP in LD with our lead SNP (*R*^2^ > 0.3) for which summary statistics were also available in the GEFOS data as a proxy. However, to avoid too weak instruments in the MR, only proxies that were associated with estradiol levels (P< 0.0001) were included in the MR. Finally, we note that any winner’s-curse bias in our effect estimates of the genetic variants associated with estradiol levels, should, on average, drive the estimates of the causal effect of estrogen on BMD towards the null.

## Results

A total of 147,690 male participants had passed our genotype and estradiol QC (Figure 1, Table 1). Of these, 134,323 had estradiol levels below detection limit (175 pmol/L), while 13,367 individuals had measured estradiol levels above, or equal to 175 pmol/L (Table 1). The number of females (N = 163,985) was slightly higher compared to males, with 37,461 having estradiol levels above detection limit, and 126,524 below. Dramatically smaller fractions of males (9.1%) and postmenopausal women (7.9%) had detectable estrogen levels (Table 1), in comparison to the fraction of premenopausal women (71.9%) with detectable estrogen levels. Number of live births, age, and BMI were significantly lower among women with detectable estradiol in both post and premenopausal strata (P<0.05).

**Table 1.** Baseline characteristics in the UKB participants included in our GWAS.

|  | Males |  | Postmenopausal Females |  | Premenopausal Females |  |
| --- | --- | --- | --- | --- | --- | --- |
|  | Above DT | Below DT | Above DT | Below DT | Above DT | Below DT |
| Participants, N (%)*** | 13499 (9.1) | 135200 (90.1) | 10090 (7.9) | 117158 (92.1) | 27672 (71.9) | 10807 (28.1) |
| Participants with all covariates, N*** | 13367 | 134323 | 9998 | 115793 | 27463 | 10731 |
| Estradiol levels (pmol/L), mean (Q1 – Q3) | 203.8 (188.8 - 230.8) | NA | 302.2 (221.2 - 475.2) | NA | 433.7 (291.7 - 686.5) | NA |
| BMI*, mean (Q1 – Q3) | **28.4 (25.2 - 30.8)** | **27.8 (25.0 - 30.0)** | **27.5 (23.6 - 30.2)** | **27.2 (23.7 - 29.8)** | **26.2 (22.7 - 28.7)** | **26.6 (22.9 - 29.2)** |
| Age*, mean (Q1 – Q3) | 57.1 (51 - 64) | 57.1 (51 - 64) | **55.0 (50 - 60)** | **60.6 (57 - 65)** | **45.9 (43 - 48)** | **47.5 (44 - 50)** |
| Smoking-Current, N (%) | 1179 (8.73) | 11424 (8.45) | **784 (7.77)** | **7445 (6.35)** | **1862 (6.73)** | **809 (7.49)** |
| Smoking-Never, N (%) | 6662 (49.3) | 65993 (48.81) | 5635 (55.85) | 67537 (57.65) | 17920 (64.76) | 6909 (63.93) |
| Smoking-Occasional, N (%) | 423 (3.1) | 4424 (3.27) | **243 (2.41)** | **1884 (1.61)** | 755 (2.73) | 281 (2.60) |
| Smoking-Previous, N (%) | 5175 (38.3) | 52905 (39.13) | 3399 (33.69) | 39842 (34.01) | 7083 (25.60) | 2788 (25.80) |
| Had hysterectomy, N (%)** | NA | NA | **4868 (48.25)** | **27841 (23.76)** | **79 (0.29)** | **76 (0.70)** |
| Hormone replacement therapy, N (never/current/previous)* | NA | NA | **3474 / 4586 / 2024** | **58112 / 4606 / 54169** | **26797 / 403 / 424** | **10166 / 157 / 463** |
| Oral contraceptives, N (never/current/previous)* | NA | NA | **1252 / 28 / 8798** | **25005 / 87 / 91874** | **2442 / 1090 / 24098** | **883 / 1476 / 8430** |
| Number of live births, median (Q1 – Q3)** | NA | NA | **1.79 (1 - 2)** | **1.9 (1 - 3)** | 1.58 (0 - 2) | 1.56 (0 - 2) |
| Age at menarche, median (Q1 – Q3)* | NA | NA | **12.62 (12 - 14)** | **12.54 (12 - 14)** | **12.68 (12 - 14)** | **12.59 (12 - 14)** |
| Age at menopause, median (Q1 – Q3)* | NA | NA | **44.62 (44 - 52)** | **46.53 (46 - 53)** | NA | NA |

DT: detection limit.
Q1: 1<sup>st</sup> quartile.
Q3: 3<sup>rd</sup> quartile.
Bold cells are significantly ( $P < 0.05$ ) associated with binary estradiol phenotype.
\*: Tested with Student's t-test.
\*\*: Tested with Fisher's exact test.
\*\*\* These are the actual number of participants analyzed in the GWAS

**Figure 1.**
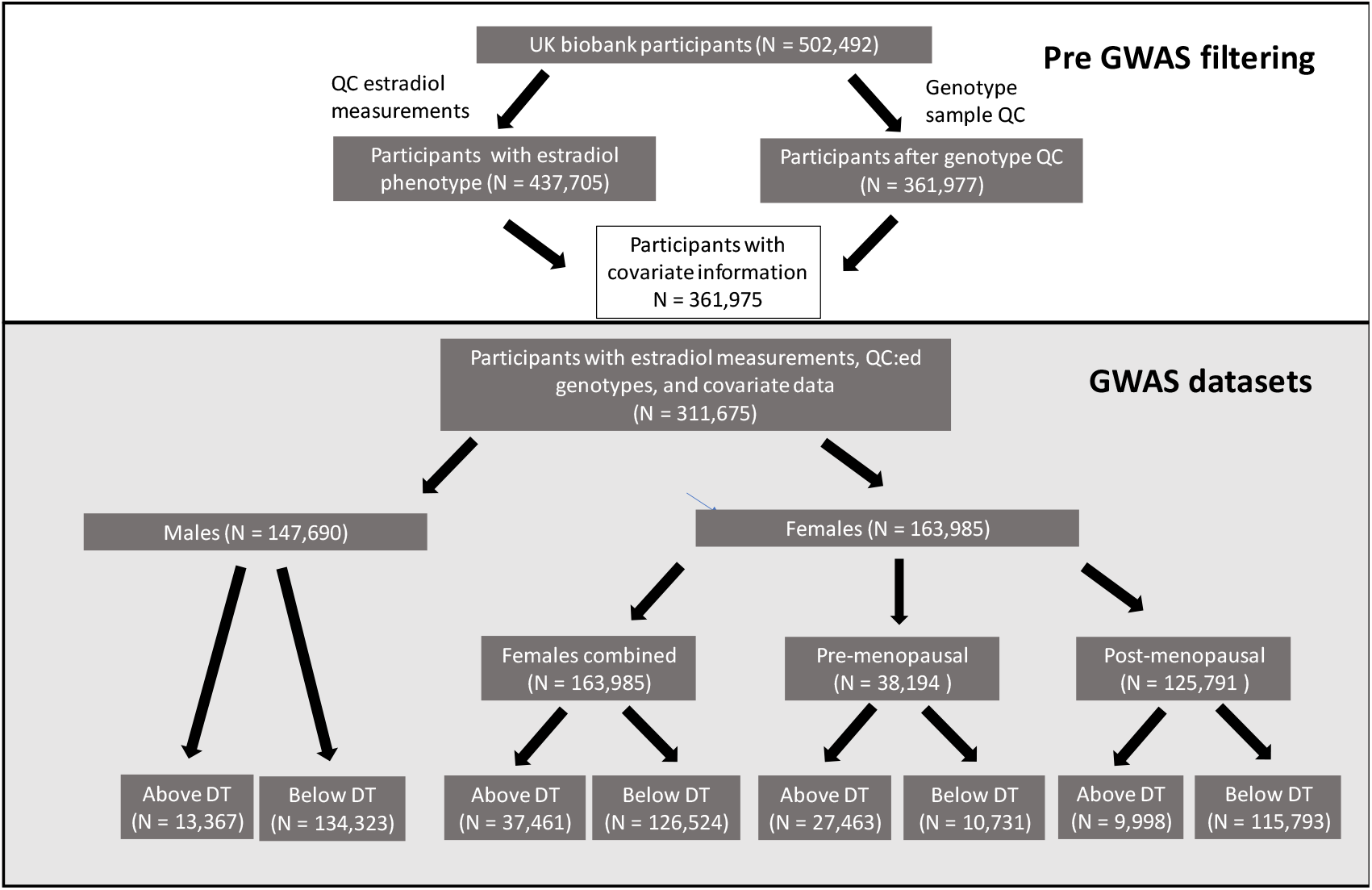
Flowchart of UK Biobank participants included in the four GWAS. QC = quality control, DT = detection limit (for estradiol measurement).

### GWAS results

We identified 15 loci, on 14 different chromosomes, to be significantly associated (P<5×10^−8^) with estradiol in males or females (Figure 2,Table 2). Of these, 13 were genome-wide significant only in males, one only in females (*MCM8*) and one in both males and females (*CYP3A7*). On chromosome 2, and 15, one conditional SNP was identified per locus, located in the vicinity to the primary lead SNPs (Table 2). Among the lead SNPs identified in males, some were located in or close to genes with a well-known role in estradiol metabolism. Among the most significant loci, we found *FAM9A* and *CYP19A1*, which have previously been identified in GWAS for estradiol in males^12,15^. One lead SNP, rs45446698, was genome-wide significant in both males and females, mapped to *CYP3A7*. It is also the most significant eQTL for *CYP3A7* in adrenal gland in GTEx (P=1.2×10^−12^), suggesting that the effect is mediated by the expression level of *CYP3A7*. The minor (G) allele increases the level of *CYP3A7* expression, but decreases the estradiol levels. There was also one lead SNP in *SHBG*, which codes for SHBG that had been linked to measurements of several other sex hormones^16^. Another lead SNP was located on the X chromosome, within the *AR* gene that encodes an androgen receptor. We also found a very strong association on chromosome 2, that overlaps with *SRD5A2*, which codes for 3-oxo-5-alpha-steroid 4-dehydrogenase 2, known to catalyze the conversion of testosterone into androgen or dihydrotestosterone^27^. Our most significant SNP is in LD (*R*^2^=0.75) with rs9282858, which is a missense variant that is predicted to be deleterious (SIFT=0.03). Here, the minor allele was associated with higher estradiol levels (Table 2). At the same locus on chromosome two, there was also one strong conditional signal, further upstream of the primary signal, closer to *MEMO1*.

**Table 2.** Results of the GWAS including conditional SNPs.

| Primary/ Conditional | Lead SNP | Chr:position | GWAS strata | Gene | Eff allele | MAF | Males |  | Females |  |
| --- | --- | --- | --- | --- | --- | --- | --- | --- | --- | --- |
|  |  |  |  |  |  |  | OR (95% CI) | P-Value | OR (95% CI) | P-Value |
| Primary | rs112881196<sup>a</sup> | 2:31982811 | males | *SRD5A2* | G | 0.039 | 1.353 (1.275 - 1.435) | 8.52E-24 | 1.106 (1.041 - 1.175)<sup>f</sup> | 0.00113 |
| Conditional | rs62142080 | 2:32182528 | males | *MEMO1/SRD5A2* | C | 0.396 | 0.927 (0.903 - 0.952) | 2.14E-08 | 0.989 (0.965 - 1.013)<sup>f</sup> | 0.384 |
| Primary | rs57102816 | 3:164506246 | males | *LINC01324* | TA | 0.257 | 1.084 (1.054 - 1.115) | 2.87E-08 | 1.008 (0.981 - 1.036)<sup>f</sup> | 5.66E-01 |
| Primary | rs7662029 | 4:69961912 | males | *UGT2B7* | G | 0.454 | 0.893 (0.871 - 0.916) | 3.61E-18 | 0.969 (0.946 - 0.992)<sup>f</sup> | 0.00945 |
| Primary | rs1073548 | 5:36012617 | males | *UGT3A1* | G | 0.126 | 1.113 (1.072 - 1.155) | 1.75E-08 | 1.052 (1.015 - 1.090) | 0.00528 |
| Primary | rs45446698<sup>b</sup> | 7:99332948 | males/females | *CYP3A7* | G | 0.043 | 0.811 (0.758 - 0.868) | 1.38E-09 | 0.813 (0.767 - 0.863) | 7.62E-12 |
| Primary | rs657152<sup>c</sup> | 9:136139265 | males | *ABO* | A | 0.340 | 0.887 (0.863 - 0.911) | 4.14E-18 | 0.955 (0.932 - 0.980)<sup>f</sup> | 0.000332 |
| Primary | rs56196860<sup>d</sup> | 12:2908330 | males | *FKBP4* | A | 0.031 | 1.263 (1.183 - 1.350) | 4.11E-12 | 0.938 (0.877 - 1.004)<sup>f</sup> | 0.0656 |
| Primary | rs11160915 | 14:106514653 | males | *IGHV3-7* | A | 0.444 | 0.849 (0.827 - 0.871) | 3.08E-35 | 0.954 (0.931 - 0.977)<sup>f</sup> | 0.000106 |
| Primary | rs28892005 | 15:51519945 | males | *CYP19A1* | A | 0.350 | 0.795 (0.774 - 0.817) | 1.01E-60 | 0.962 (0.939 - 0.986)<sup>f</sup> | 0.00227 |
| Conditional | rs3751591 | 15:51606710 | males | *CYP19A1* | G | 0.167 | 1.125 (1.088 - 1.163) | 3.22E-12 | 1.004 (0.972 - 1.036)<sup>f</sup> | 0.811789 |
| Primary | rs62059839 | 17:7533015 | males | *SHBG* | T | 0.262 | 1.098 (1.067 - 1.130) | 8.53E-11 | 1.044 (1.016 - 1.072) | 0.00184 |
| Primary | rs113047993 | 18:20585399 | males | *RBBP8* | T | 0.068 | 0.858 (0.813 - 0.904) | 1.47E-08 | 0.977 (0.932 - 1.024)<sup>f</sup> | 0.337 |
| Primary | rs62129966 | 19:48374950 | males | *SULT2A1* | A | 0.164 | 1.113 (1.077 - 1.150) | 2.18E-10 | 1.012 (0.981 - 1.045)<sup>f</sup> | 0.456 |
| Primary | rs16991615 | 20:5948227 | females | *MCM8* | A | 0.065 | 1.043 (0.992 - 1.097) | 0.101 | 1.139 (1.087 - 1.193) | 4.67E-08 |
| Primary | rs5933688<sup>e</sup> | X:8880680 | males | *FAM9A* | G | 0.275 | 1.118 (1.097 - 1.140) | 7.49E-30 | 1.013 (0.986 - 1.040)<sup>f</sup> | 0.348 |
| Primary | rs114255570 | X:67005508 | males | *AR* | A | 0.077 | 0.899 (0.867 - 0.931) | 5.6E-09 | 1.002 (0.959 - 1.047)<sup>f</sup> | 0.919 |

<sup>a</sup> LD with rs9282858, which is missense variant in *SRD5A2* (SIFT=0.03 deleterious, PolyPhen2=0.337 benign).<sup>7</sup>
<sup>b</sup> eQTL for CYP3A7.
<sup>c</sup> rs657152 tags the O blood group.
<sup>d</sup> rs56196860 is a missense variant in FKBP4.
<sup>e</sup> rs16991615 is a missense variant in MCM8.
<sup>f</sup> Significant (P<0.05) difference in OR between males and females.

**Figure 2.**
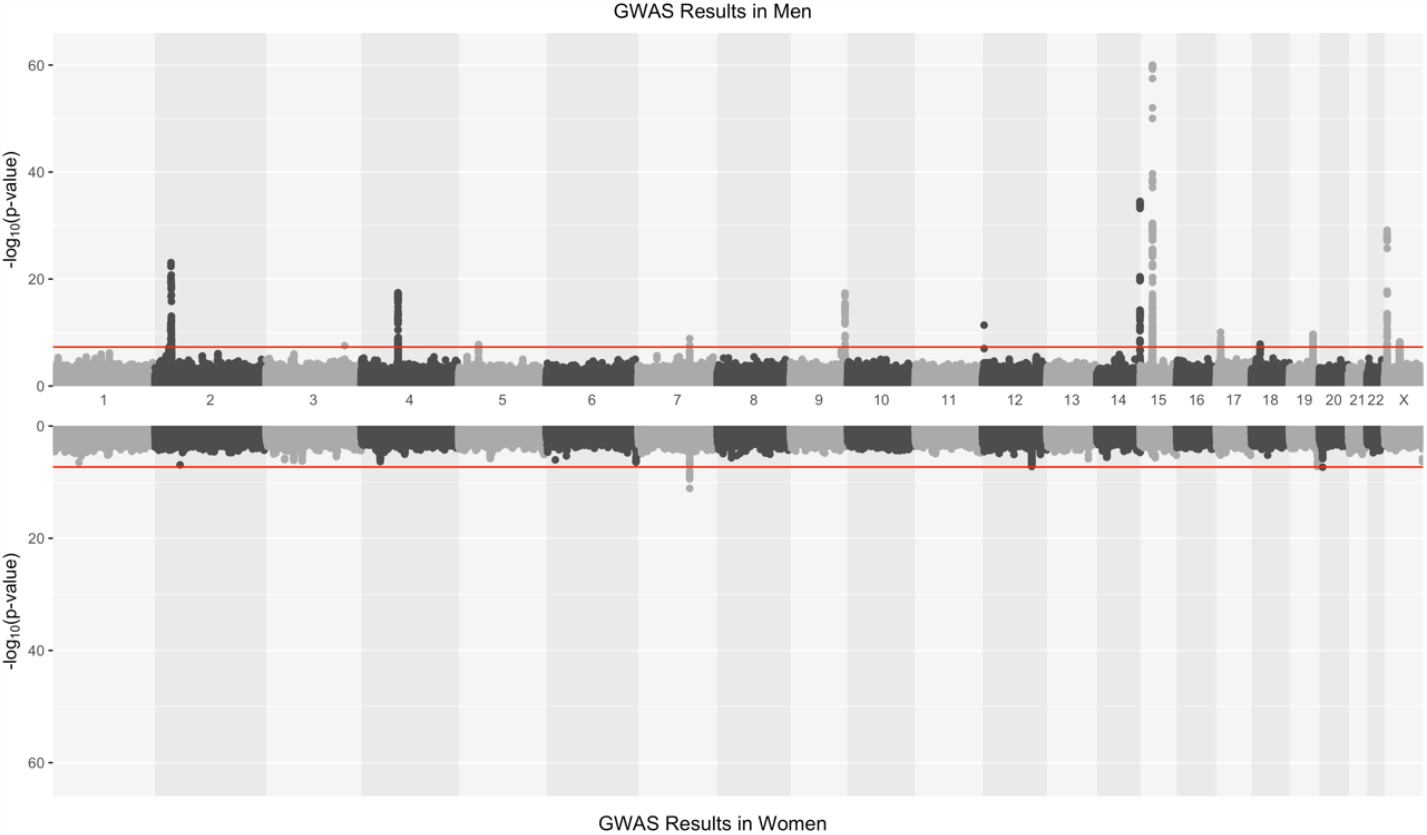
Hudson plot for the estradiol GWAS in males (upper) and females (lower). The red horizontal line indicated the genome-wide significance threshold (5×10^−8^).

There was also an association to rs657152 in *ABO*, and SNP that is in LD (*R*^2^=0.98) with rs8176719, which tags the O blood group^28^. The A allele of rs657152 (Table 2) tags the non-O blood groups and we found rs657152-A to be associated with lower levels of estradiol, indicating that individuals with a non-O blood group have lower levels of estradiol compared to individuals with the O blood group. Two associations on chromosome 4 and 5 respectively, mapped to *UGT2B7* (rs7662029) and *UGT3A1* (rs1073548). UGT3A1 and UGT2B7 are glucuronosyltransferases, which are enzymes that catalyze the glucosidation of lipophilic chemicals, facilitating their elimination^29^. Another lead SNP mapped to *SULT2A1*, which encodes a sulfotransferase which is believed to be important for sex-steroid biosynthesis by sulfating hydroxylated steroid hormones and bile acids^30^. Another lead SNP on chromosome 14 is a missense variant in *FKBP4*, which encodes the protein FKBP52 that has been suggested to be involved in the formation of steroid hormone nuclear receptors that regulate hormone levels^31^.

The remaining lead SNPs from the GWAS in males were mapped to loci with less clear links to estradiol metabolism. These include one lead SNP (and one conditional SNP) in a locus with a cluster of genes, including *IGHV3-7* and *IGHV6-1*, that encodes parts of immunoglobulin heavy chains that participates in the antigen recognition. Another lead SNP is located in *RBBP8* which encodes Retinoblastoma-binding protein 8 (RBBP8, CtIP), an endonuclease involved in the repair of double-stranded DNA breaks through homologous recombination as well as regulation of G2/M cell-cycle checkpoints as part of the BRCA1-RBBP8 complex^32^. Variation in *RBBP8* genotype and regulation has been reported in multiple types of cancer^33,34^. A third SNP was a missense variant in *LINC01324*, a lncRNA which has been established as a predictor for melanoma progression^35^.

The lead SNP that was only significant in females (rs16991615, Table 2), is a missense variant located in *MCM8. MCM8* codes for a DNA helicase, which is involved in the repair of double-stranded DNA breaks and recombination^36^. Even though very few genome-wide significant SNPs were identified in women, eight out of 14 of the loci that were genome-wide significant in males, were at least nominally associated in females as well (P < 0.05), and with the same direction of effect (Figure 3). Especially the genes with functions related to hormone metabolism; *UGT2B7, UGT3A1, CYP3A7, CYP19A1, SRD5A2*, and *SHBG* replicated in females, but also *IGHV3-7* and *ABO*.

**Figure 3.**
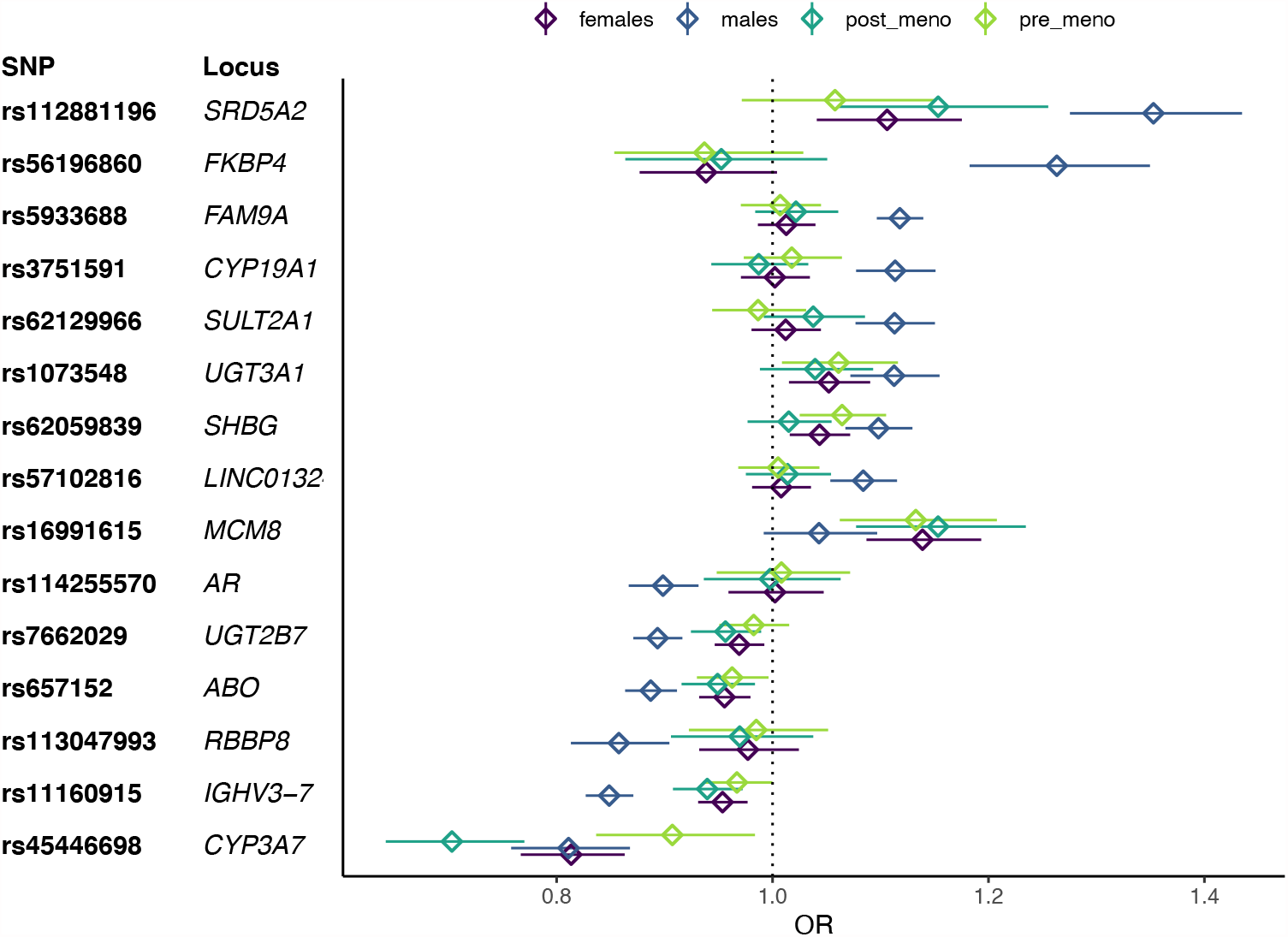
OR and 95% CIs for the lead SNPs in the male and female strata, as well as the two female subgroups: pre- and post-menopause. The lead SNP for each locus that reached genome-wide significance in males or females in the primary analyses are included. Non-overlapping confidence intervals typically indicate a significant difference in ORs well below p = 0.05.

### Sensitivity analysis

Even though the primary aim of the sensitivity analyses was to further verify the associations from the primary GWAS, we did perform GWAS with the different sets of covariates/strata in the sensitivity analyses as well. However, no additional genome-wide significant locus was identified. The effects of the findings from the primary analyses changed somewhat. When stratifying females into pre- and post-menopause there was one locus, *CYP3A7*, with a significant difference in the OR between the two strata (Supplementary Table S1^37^, Figure 3). For *CYP3A7*, the largest effect was seen in postmenopausal women OR = 0.703 (0.642 - 0.770), followed by males OR= 0.811 (0.758 - 0.868), and then premenopausal OR = 0.907 (0.837 - 0.984).

After removing all participants that had a prior cancer diagnosis at the time of assessment, there was no significant difference in OR compared to the primary GWAS, and all lead SNPs were still genome-wide significant, indicating that a prior cancer diagnosis did not influence our primary results (Supplementary Table S1^37^). However, when adjusting the estradiol GWAS for testosterone and SHBG levels, the lead SNPs at *SHBG* and *FKBP4* were no longer genome-wide significant in males, and the corresponding effects were significantly lower than the previous signals. Also, for *UGT3A1* and *AR*, the P-values were below the threshold for genome-wide significance for estradiol when adjusting for testosterone and SHBG levels, but the ORs were not significantly different from the primary analyses. This indicates that the signals at *SHBG* and *FKBP4* might have been completely driven by SHBG and testosterone levels, whereas the other loci are more likely to be estradiol specific loci. We observed the same effect for the female-specific locus *MCM8*. There was no significant difference in OR between the primary GWAS in postmenopausal women with and without adjustment for age at menopause, suggesting that the results in females are not strongly confounded by age at menopause as has been discussed previously^16^.

### GWAS overlap with estradiol and related phenotypes

We did a systematic check for overlap with previous GWAS results that were available in the GWAS catalog (Supplementary Table S2^37^). For lead SNPs at *UGT3A1, LINC01324* and *RBBP8*, we found no overlap with previous GWAS signals. Several SNPs at the *AR* locus have previously been associated with male pattern baldness^38^, but none of those are in strong LD with our lead SNP (*R*^2^<0.3). The lead SNP at the other ten loci, all overlapped with at least one previous GWAS. There were several overlapping signals with phenotypes that are related to sex hormones. For example, the lead SNPs at *SRD5A2* and *FAM9A* overlap with male pattern baldness, as well. The lead SNPs at *CYP3A7* and *SULT2A1* overlap with GWAS-hits for several sex hormones and other metabolites, *UGT2B7* overlaps with serum and urinary metabolites, while *SHBG* overlaps with testosterone levels. Our lead SNPs in *FAM9A, CYP19A1*, and *CYP3A7* all overlap with previous GWAS for BMD. Other loci overlap with GWAS for more distal traits. For example, the lead SNP in *ABO* overlaps with results from over 100 GWAS, with associations to metabolites, liver enzymes and proteins as well as cardiovascular diseases, cancer, and type 2 diabetes. The lead SNP at *FKBP4* overlaps with waist to hip ratio and lung function. *MCM8*, the female specific locus, overlaps with traits related to age at menopause. This agrees partly with our sensitivity analyses where the OR decreased slightly from OR = 1.139 (1.087 – 1.193) in the primary GWAS to OR = 1.126 (1.035 – 1.225) when adjusted for age at menopause. Still the OR was significantly different from unity in the sensitivity analyses suggesting that the locus is probably associated with both age at menopause and estradiol levels.

### Mendelian randomization

A total of 209,043 participants (96,432 males and 112,611 females) had BMD measurements and covariate information available, and were included in the MR analysis. Mean value for T-scores in males was −0.085, and for females −0.58. The average T-scores below zero agrees with the average age in UKB participant being higher than the general population, and thereby the BMD being lower compared to the mean BMD of respective sex. The instrument variables for the MR analyses were selected from the lead SNPs in the primary GWAS for males and females, respectively. For males, all significant lead SNPs and conditional SNPs were selected, resulting in 16 instruments (Table 3). The estimated effects on estradiol level (Supplementary Table S3^37^) were taken from the primary GWAS for all instruments, also for the SNPs that were identified in the conditional analyses. Hence, the estimates for the conditional SNPs differ slightly from the ones in Table 2. Using GSMR, three instruments were identified as pleiotropic and were excluded from the primary analysis in males (Supplementary Table S3^37^). We identified a causal effect of high estradiol levels on BMD in males (Table 3, Figure 4a). As estradiol was analyzed as a binary phenotype, the unit for the estimate from the MR can be interpreted as standard units increase in T-score associated with having estradiol levels above detection limit. Similar MR results were obtained using weighted median and MR-Egger, which do not remove pleiotropic SNPs. The MR-Egger intercept was not significantly different from zero, indicating that these results are not strongly influenced by directional pleiotropy.

**Table 3.** Results of MR analyses.

|  | Males<br>(one-sample MR: UKB) |  | Males<br>(two-sample MR:<br>UKB - GEFOS) |  | Females<br>(one-sample MR: UKB) |  | Females<br>(two-sample MR:<br>UKB - GEFOS) |  | Females, with SNPs from males<br>(one-sample MR: UKB) |  |
| --- | --- | --- | --- | --- | --- | --- | --- | --- | --- | --- |
| MR method | Estimate | P-value | Estimate | P-value | Estimate | P-value | Estimate | P-value | Estimate | P-value |
| GSMR | 9.89E-02 | 1.57E-11 | 1.85E-01 | 1.20E-04 | 3.27E-02 | 7.48E-06 | 4.08E-01 | 3.49E-05 | 1.64E-01 | 4.97E-06 |
| Weighted median | 6.61E-02 | 2.82E-02 | 2.62E-01 | 1.01E-04 | 7.31E-02 | 1.26E-01 | 4.48E-01 | 1.25E-04 | 1.14E-01 | 1.49E-01 |
| IVW | 1.48E-01 | 5.49E-05 | 1.84E-01 | 1.06E-03 | 1.45E-01 | 9.98E-02 | 3.05E-01 | 6.11E-02 | 1.64E-01 | 1.93E-01 |
| MR-Egger | 1.42E-01 | 2.23E-01 | 4.51E-01 | 1.67E-02 | 4.81E-01 | 2.56E-02 | 5.84E-01 | 4.98E-01 | 3.08E-01 | 2.73E-01 |
| MR-Egger intercept | 8.87E-04 | 6.95E-01 | -4.02E-02 | 8.36E-02 | -3.63E-02 | 1.71E-01 | -2.56E-02 | 7.23E-01 | -1.09E-02 | 5.17E-01 |

**Figure 4.**
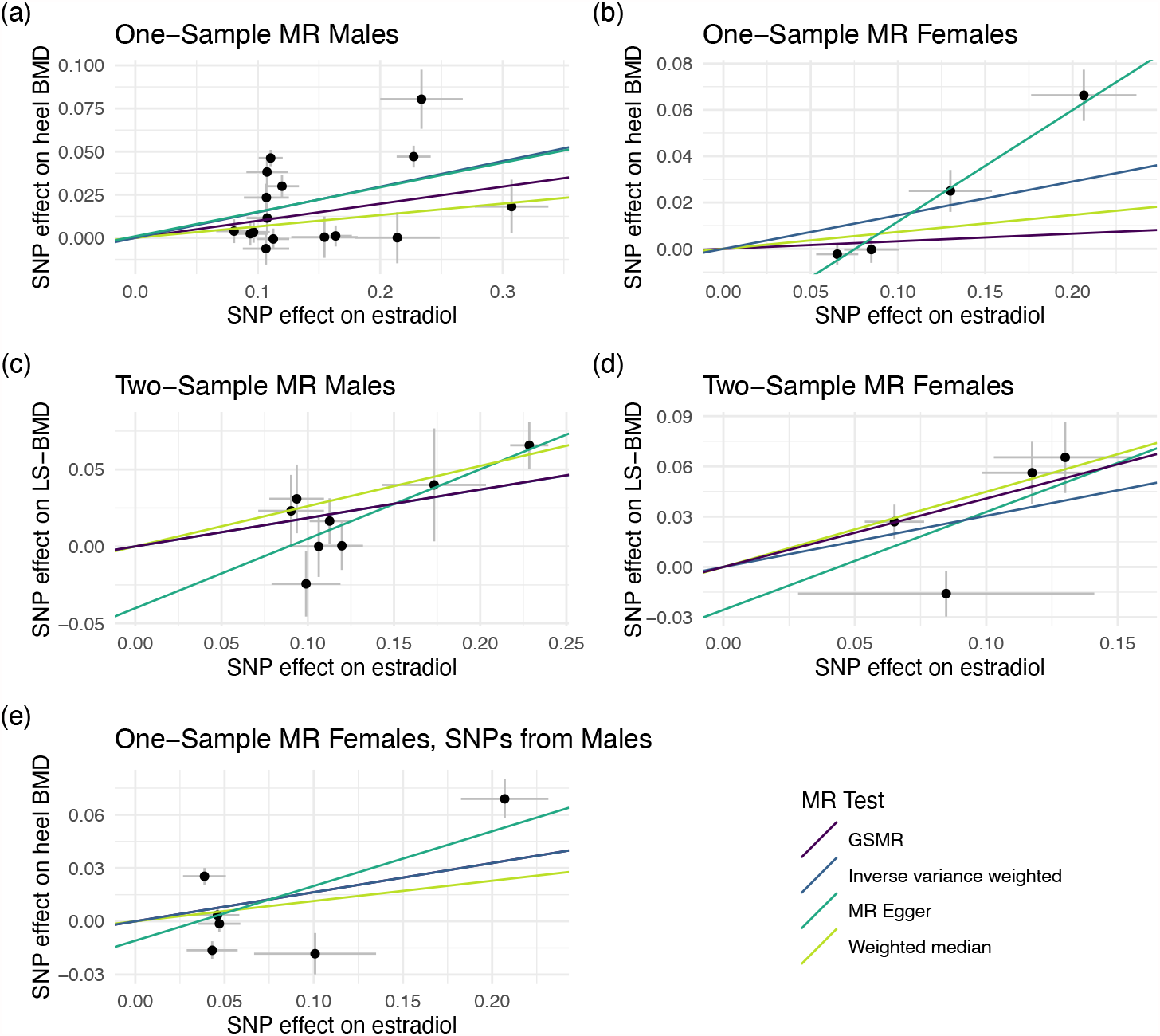
Mendelian randomization analyses to estimate the causal effect of estradiol on bone mineral density (BMD). (a) One-sample MR in UKB males, (b) One-sample MR in UKB females. (c) Two-sample MR with GEFOS males, and (d) Two-sample MR in GEFOS females. (e) One-Sample MR analysis with estradiol instruments selected in the UKB males, and MR performed in UKB females.

To get a reasonable number of instruments to be used for the MR analysis in females, we used a less strict P-value cutoff (P<1×10^−7^), resulting in a total of 4 instrumental variables (Supplementary Table S3^37^) that were included in the analysis. In the GSMR analyses, the HEIDI-outlier flag was disabled in females, as there were fewer (<10) SNPs than recommended for this approach to work properly^24^. We identified a causal effect of high estradiol levels on BMD also in women (β=0.03, P=7.48×10^−6^, Figure 4b). All three alternative MR methods showed the same direction of effect, although the null hypothesis of no effect could not be rejected, and the MR egger intercept was not significantly different from zero (Table 3).

To further validate the MR results we also did a two-sample MR for males and females respectively using the instruments selected for estradiol in UKB and the GWAS summary statistics from the BMD GWAS in GEFOS^26^. A slightly different set of instrumental variables (Supplementary Table S3^37^) had to be selected, since considerably fewer SNPs were analyzed in GEFOS due to their use of the HapMap reference panel for imputations. We included 8 instruments for the replication in males, and four in females. The MR estimates were in the same direction as the one-sample MR (Figure 4c, d), but with a larger effect in both males (β=0.18, P=0.00012) and females (β=0.41, P=3.49×10^−5^)

Previous concerns that age at menopause influences estradiol levels, could potentially render some of our selected instruments invalid since being affected by confounding (correlated horizontal) pleiotropy. We therefore also performed a third set of MR-analyses in females.

Here, we selected the instruments for estradiol from the GWAS in males, which should not be influenced by age at menopause. To reduce the risk for male-specific effects on estradiol levels, we filtered for SNPs that were significant also in females, after adjusting for the 16 instruments tested (P<0.0031), resulting in six instruments and used the estradiol estimates for females in UKB (Supplementary Table S3^37^). Using the GSMR approach, we could identify a significant effect on BMD in females, similar to the effect estimated in males (β=0.16, P=4.97×10^−6^ (Table 3, Figure 4e). These results indicate that the effect in females is not driven by age at menopause.

## Discussion

We have presented results from a large GWAS for estradiol levels in males and females. Many of the significant SNPs are annotated to genes with functions that are biologically relevant to estrogen metabolism. For example, by converting and transporting androgen and endogens, synthesis or elimination of steroid hormones. In males, we identified 14 loci, to be associated with serum estradiol levels, and another two independent SNPs on chromosome 2 and 15. In females, we identified two significant loci. One locus, within the *CYP3A7* gene, overlaps with our findings in males, but the other locus, mapping to *MCM8*, was specific to females. However, a majority (eight) of the male loci, was at least nominally significant in females, even if the estimated effect size was lower. These results suggest that genetic effects on estradiol levels might differ between males and females. However, in premenopausal women, estradiol levels vary during the menstrual cycle, limiting our ability to estimate the genetic effects on estradiol in females. In postmenopausal women, on the other hand, estradiol levels drop rapidly, and it has previously been suggested that the estradiol levels in females are more likely to be determined by the number of years since menopause^39^. For those reasons, we performed sensitivity analyses, stratifying females by menopausal status, and also adjusting for age at menopause in the postmenopausal strata. The only locus being significantly different between pre- and postmenopausal women was *CYP3A7*. The largest effect was seen in postmenopausal women, followed by males, and then premenopausal women. *CYP3A7* encodes cytochrome P450 CYP3A7, which metabolizes dehydroepiandrosterone (DHEA), a precursor of both androgen and estrogen synthesis, and shows catalytic activity for conversion of estrone to hydroxyestrogens^40,41^. As much as 75% of the circulating estrogens in premenopausal women and 50% of circulating androgens in men are known to be derived from DHEA. After menopause, DHEA is thought to be the major precursor of androgens and estrogens in females^42^. Interestingly, the effect of the *CYP3A7* SNP disappeared when adjusting for SHBG and testosterone levels in females, but the effect was unaffected in males. However, *CYP3A7* loses genome-wide significance when adjusting for testosterone levels. This suggests that *CYP3A7* might have different functions in males and in females, as well as in pre-and in postmenopausal women. In postmenopausal women *CYP3A7* appears to mainly influence estradiol levels by regulating the amount of available testosterone as a precursor for estrogen.

Several other loci, with previous links to estrogen metabolism were also identified. *SULT2A1* encodes a sulfotransferase which are active on hydroxylated steroid hormones and bile acids^30^. Sulfotransferase 2A1, encoded by *SULT2A1*, mediates the sulfation of a wide range of steroids and sterols, including DHEA, a precursor of estrogen and testosterone. *SULT2A1* was only associated with estradiol in males, and this association was similar (slightly stronger) when adjusting for testosterone and SHBG levels. Two of the most well-known loci, that has previously also been associated with estradiol in males in GWAS, are *FAM9A* and *CYP19A1*^12,15^. Aromatase, encoded by *CYP19A1*, is a key enzyme in the synthesis of estradiol and estrone from testosterone and androstenedione, respectively^43^. The association to *CYP19A1* was at least nominally significant, in both the pre- and postmenopausal women, as well as in all sensitivity analyses. The P-values were even numerically slightly lower when adjusting for SHBG and testosterone, which agrees with the function of the protein. *FAM9A* is exclusively expressed in testis^59^ and it is therefore not surprising that there was no association to *FAM9A* in females. Previously, *FAM9A* has also been associated with testosterone levels^15^, and our analyses indicates that the association was partly driven by associations to testosterone or SHBG levels. We could therefore conclude that the SNP at *FAM9A* is likely to affect both testosterone and estrogen metabolism. This signal also overlaps with a GWAS signal for male pattern baldness^38^, which further supports the importance of this locus. However, *FAM9A*, encodes the protein FAM9A, that has a rather unknown function. Also, the SNP is located between *FAM9A* and *FAM9B* and the annotation of this locus should therefore be interpreted with care.

We identified a strong association between estradiol levels and the *SHBG* locus. The *SHBG* gene encodes SHBG that has been linked to measurements of several other sex hormones previously^16^. SHBG is a glycoprotein that binds to sex hormones, including testosterone and estrogen, in the bloodstream, and thereby regulates the amount of bioavailable hormones^44^. Only 1–2% of sex hormones are unbound and therefore bioavailable^45–47^. The association to SHBG in males, completely disappeared after adjusting for SHBG levels. In females, there was a nominally significant association to SHBG in the primary GWAS, with the same direction of effect as in males. Interestingly, after adjusting for SHBG and testosterone levels, there was still an association (P=0.00033), but the effect was in the opposite direction. Similarly, an opposite effect between males and females has previously been seen for total testosterone, where there is a negative genetic correlation between total testosterone and SHBG in females, but a positive correlation in males^16^. This clearly illustrates the heterogenous effects between males and females for sex hormones. We also identified two associations to the X chromosome in males, one mapping to the *AR* gene, which encodes the androgen receptor. In mammals, the action of androgens acts through this single androgen receptor. In different tissues, the androgen receptor is activated by testosterone, or testosterone is first converted into dihydrotestosterone which is then the activating molecule^48^. Therefore, the androgen receptor plays an important role in sex differentiation, mainly through regulating gene expression^49^. Interestingly no association (P>0.05) was seen in females which might be due to the much lower levels of androgens in females. We also found an association to *SRD5A2* which encodes 3-oxo-5-α-steroid 4-dehydrogenase 2 and is located on chromosome 2. This enzyme catalyzes the conversion of testosterone to 5-α-dihydrogentestosterone^27^. Thus, the variant in *SRD5A2* might influence estradiol levels in men by affecting the amount of testosterone available for conversion to estradiol. The SNP in *SRD5A2* is nominally significant in females, an association that is more pronounced in postmenopausal women. There is also a conditional signal in males, 200 kb upstream, close to *MEMO1* which encodes the mediator of ErbB2-driven cell motility 1 (MEMO1). In previous studies, it has been shown that MEMO1 regulates the extranuclear function of the estradiol receptor^50^, which is a possible link to estradiol levels. It is therefore possible that the GWAS signal on chromosome two, indeed represent associations with two different genes. Another interesting association is with *FKBP4*, which encodes FKBP52 (FK506-binding protein 4). FKBP52 has been show to regulate the androgen, glucocorticoid and progesterone receptor signaling pathways^31^, by playing an important role in the formation of steroid hormone nuclear receptors^51^. *FKBP4* has been shown to regulate androgen receptor transactivation activity and also been linked to testosterone levels^52^. When adjusting for testosterone and SHBG levels, the *FKBP4* association was weaker, and no longer significant.

Two loci, mapping to *UGT2B7* and *UGT3A1* were also identified. These two loci encode UDP-glycosyltransferases, which are responsible for the addition of sugars to a broad range of lipophilic molecules to improve the water solubility and thereby play a major role in elimination of potentially toxic xenobiotics and endogenous compounds. Both UGT3A1 and UGT2B7 have been shown to metabolize estrogens by glucuronidation, i.e., the transferring of glucuronic acid to the metabolites^29,53,54^. This suggests that UDP-glycosyltransferases might play a major role in estradiol metabolisms. Also, *ABO* encodes a glycosyltransferase that is known to modify carbohydrates on the red blood cell antigens. *ABO* is the gene that is responsible for the ABO blood groups^28^. The effect allele of our study (the A allele) is in LD with the non-O blood groups, indicating that men with a non-O blood type have lower estrogen levels, or the other way around, men with blood group O have higher levels. Since *ABO* and its histo-blood group properties have been linked to a vast number of diseases and functions, it is not unlikely that our finding is indicative of an association between ABO antigens and estradiol levels. The most well-known association for ABO, is that individuals with the O blood group have lower risk of cardiovascular diseases^55^, probably due to lower coagulation activity. There are very limited findings of a clear link between ABO and estrogen levels. However, one study found an interaction between ABO and HRT in relation to coagulation activity^56^. ABO antigens are present on most epithelial and endothelial cells as well as on T cells, B cells and platelets and are also detectable in most body fluids such as saliva^57^. Recently, ABO was linked to Covid-19, where the susceptibility to SARS-CoV-2 has been suggested to be explained by modulation of sialic acid-containing receptors distributed on host cell surface^58^. Covid-19 has a higher mortality in males, which has been suggested to be linked to their lower estrogen levels^59,60^. We also identified several loci with less clear links to estrogen metabolism including *LINC01324, IGHV3-7, RBBP8*, and *MCM8. MCM8* is the female-specific locus, where the lead SNP is a missense variant. Genetic variants in *MCM8* have previously been associated with premature ovarian failure^61,62^, a condition that is characterized by low estrogen levels^51^. According to GTEx, *MCM8* is highly expressed in both female and male reproductive organs, despite only being significant in the female GWAS cohort, which further support this locus as being involved in sex hormone metabolism.

We also performed MR analysis and estimated a causal effect of estradiol on BMD in males, and to our knowledge, for the first time also in females. Compared to previous studies in males, with not more than five SNPs as instruments for estradiol^17^, we included as many as 16 SNPs in males and four in females. Interestingly, the effect estimate was higher for females than males in the two-sample MR, which could agree with the fact that BMD decreases more rapidly in females by age. Estrogen can be used as a preventative treatment in postmenopausal women with low BMD^63,64^, to prevent osteoporosis, and our results were therefore not surprising. However, one concern that has been raised previously^16^, is that estradiol levels are driven by number of years till/from menopause. To reduce potential bias in our results due to instruments being associated with age at menopause, we applied a second approach where we used instruments from males, to minimize our instruments’ association to age at menopause. Using these instruments, we could replicate the MR results in women which suggests that age at menopause is not dramatically influencing our results.

One of the major strengths of this study is the large cohort from UKB with estradiol measurements available. However, only a subset of the participants had estradiol levels above detection limit. To increase the number of participants to analyze, we created a binary phenotype. Since normal levels of estradiol in males and postmenopausal women have a range below or just above the detection limit, we were not able to capture the full spectrum of individual variation in the cohort. Also, very few loci were identified in females. This could be due to hormone levels fluctuating during the menstrual cycle lowering the power to identify true genetic effects behind estradiol levels in premenopausal women. However, we also conducted a GWAS including only women that had entered menopause, which should give a more stable phenotype. However, no additional significant GWAS hits were identified, even if the number of participants in the GWAS was comparable to the GWAS in males, and the range of estradiol levels was comparable. This suggest that there is an underlying difference in genetic effects between sexes.

In summary, we have identified biologically relevant genetic loci associated with variation in estradiol levels in males and females and found differences in genetic effects between sexes. In addition, we used an MR approach to show that estradiol levels have a causal effect on good bone health in both males in females, confirming that estrogen replacement therapy may be a good treatment to prevent osteoporosis and prevent fractures in elderly of both sexes. Our findings support previous research on the synthesis of estradiol and its effects on bone mineral density, as well as provide new insights into the genetic components of estrogen regulation and metabolism.

## Data Availability

The data used for this study is available for bona fide researchers from the UK Biobank Resource (http://www.ukbiobank.ac.uk/about-biobank-uk/), and can be accessed by an application to the UK Biobank. GEFOS data included in this study can be downloaded from http://www.gefos.org.

https://doi.org/10.5281/zenodo.4575527

## Acknowledgements

This research was conducted using the UKB Resource under applicationnumber 41143, following the restrictions on data availability set up by the UKB. We acknowledge all participants and staff involved in UKB for their valuable contribution. The computations were performed on resources provided by SNIC through Uppsala Multidisciplinary Center for Advanced Computational Science (UPPMAX) under project sens2017538. The research was funded by the Swedish Research Council (Å.J), and the cancer (Å.J), the brain (Å.J), the heart and lung (Å.J.), the Å Wiberg (W.E.E), M Borgström (W.E.E), Hedströms, K och O F (W.E.E), A and M Rudbergs (W.E.E) foundations.

## Author contributions

W.E.E and Å.J designed the study; E.B, W.E.E, D.S performed the data analysis; D.S generated the figures; W.E.E, D.S and Å.J wrote the manuscript; W.E.E, T.K, D.S, E.B, Å.J, J.H interpreted the data, contributed to and reviewed the manuscript.

